# Family responsibilities and mental health of kindergarten educators during the first COVID-19 pandemic lockdown in Ontario, Canada

**DOI:** 10.1101/2021.05.11.21257057

**Authors:** Natalie Spadafora, Caroline Reid-Westoby, Molly Pottruff, Magdalena Janus

## Abstract

The present study, conducted during the first wave of the COVID-19 pandemic in Ontario, Canada, addressed the association between family responsibilities and mental health (depression and anxiety) among kindergarten educators. Participants comprised 1,790 (97.9% female) kindergarten educators (73.6% kindergarten teachers; 26.4% early childhood educators) across Ontario. Results revealed that educators were more likely to report moderate levels of depressive symptoms if they had the responsibility of caring for their own children, and more likely to report moderate levels of depressive and anxious symptoms if they had the responsibility of caring for an older adult. Theoretical and practical implications are discussed.

## 1. Introduction

The COVID-19 pandemic and related public health restrictions placed on society have dramatically affected many areas of everyday life. School closures, for example, impact students, families, as well as teachers and educators (e.g., Müller & Goldenberg, 2020). As a result of the pandemic-related school closures, elementary school education shifted to distance learning in the spring of 2020 using online platforms, with very little preparation time (Jeffords, 2020). Since the beginning of the school closures, many teachers acknowledged the burden of adapting lessons from a physical classroom to an online format and viewed this arrangement as exhausting and not sustainable (Turner et al., 2020). Teachers were faced with a myriad of new challenges including reduced communication with students and their families, increased technological barriers, and students opting out of learning altogether (e.g., Fauzi & Khusuma, 2020). Some teachers even supported fellow colleagues who had less experience with digital platforms to help them set up their online classes, which led to overworking themselves (Lieu, 2020). In addition to this, many teachers had increased family responsibilities as a result of the COVID-19 pandemic and its associated isolation and quarantine rules. For example, educators with school-aged children were suddenly expected to teach and interact with their students from a distance, while also caring for and overseeing the learning of their own children, resulting in a great deal of added stress (Košir et al., 2020). Further, some teachers were also responsible for the care of an older adult, such as running errands and preparing meals, all the while trying to balance isolation measures, which also may have increased stress (Kent et al., 2020). However, the extent of the association between caregiving responsibilities and educators’ mental health during the COVID-19 pandemic is not yet known.

Previous research, conducted prior to the pandemic, has long established the link between the burden of caregiving and poor mental health. For example, a meta-analysis by Pinquart and Sörensen (2003) found that having caregiving responsibilities was positively associated with depressive moods and that caregiving-related stress was more strongly associated with poor mental health compared to other life stressors. Research examining caregivers of ill family members in Taiwan demonstrated that the burden of caregiving was associated with poorer mental and physical health (Chang et al., 2010). Another study by Sales and colleagues (2014) found that the relationship between child behavioral problems and mothers’ mental health was mediated by the caregiving strain of the mother. While existing literature has demonstrated an association between caregiving and mental health, much of the literature to date has focused on the burden of caregiving for a child with behavioral issues, special needs, or other health issues (e.g., Baykal et al., 2019; Meltzer et al., 2011; Rosenzweig et al., 2008). Further, research has previously examined challenges faced by working parents, particularly females, reporting that caregiving responsibilities can negatively impact their quality of life, including lack of time to spend with their friends (Lahaie et al., 2013). As the COVID-19 pandemic necessitated school closures, the burden of parenting increased for all parents, creating unique challenges for working parents in particular.

### 1.1. Caring for Family Members During the COVID-19 Pandemic

Recent research examining the effects of the COVID-19 pandemic has highlighted high levels of caregiver stress and burnout (e.g., Brown et al., 2020; Russell et al., 2020). Fulfilling the dual role of working and caring for their own children, and potentially supporting their learning, has been an added stressor for parents working from home during the pandemic. Research has suggested that the increased burden of childcare during the COVID-19 pandemic has been associated with parent perceptions of their child’s heightened stress as well as their own poorer mental health (Russell et al., 2020). In addition, higher parental stress associated with the COVID-19 pandemic was also associated with higher levels of anxiety and depressive symptoms (Brown et al., 2020).

Research conducted in June 2020 in the United States found that 27% of parents of children under 18 years reported a decline in their own mental health since the start of the pandemic (Patrick et al., 2020). In the same study, 1 in 10 families reported worsening of parents’ mental health and of the behavior of their child, with almost 50% reporting loss of regular childcare. On the other hand, another recent study found that taking care of children following pandemic-related closures of childcare centres and schools benefitted the emotional well-being of parents as it was associated with higher positive affect and lower negative feelings (Lades et al., 2020). However, this study also found an increase in negative affect for parents who were responsible for their children’s schooling within the home. Košir et al. (2020) conducted a study on teachers and school counsellors working in Slovenian elementary schools during the COVID-19 school closures and found that educators who reported caring for their own preschool or younger elementary school-aged children at home expressed experiencing higher levels of stress related to distance teaching. Recent data examining the impact of the pandemic on Canadians from May to November 2020 also found that Canadian adults with children under 18 years reported feeling more anxious than those who did not (CAMH, 2020).

Working professionals who have the responsibility of caring for older adults may also face increased stress and poorer mental health outcomes during the pandemic. Kent et al. (2020) highlighted the increasingly important role of family caregivers with regards to caring for elderly family members during the pandemic, given the additional medical challenges during this time. Kent et al.’s study highlighted an increased burden on adult caregivers, as balancing caregiving with social isolation measures could be complex and stressful. Moreover, research focused on caregiving for elderly adults with dementia found that having this responsibility was associated with higher levels of depression and anxiety during the COVID-19 pandemic quarantine (Altieri & Santagelo, 2021). Overall, given the added stress of the pandemic and related lockdowns on both the teaching profession and family caregiving, it is important to understand whether educators who have the added responsibility of caring for children or older adults are facing increased mental health challenges as a result of the COVID-19 pandemic.

### 1.2. Why Teacher Mental Health Matters in Times of School Closures

Even in non-pandemic situations, teacher stress and feelings of burnout are associated with their own poor psychological well-being and more negative emotions towards students (Burić et al. 2019) and with elevated stress of the children in their elementary school class (Oberle & Schonert-Reichl, 2016). Previous research also found that teachers felt emotionally burnt out when they did not feel respected and felt accomplishment burnout when there was a lack of sociability and personalization (Hastings & Bham, 2003). As we anticipate the general levels of stress and burnout during the COVID-19 pandemic to increase (e.g., Sokal et al., 2020), it is particularly important to understand teacher well-being during this time.

Based on existing literature on the impact of pandemics, it is evident that teachers play a pivotal role in helping children and their parents navigate school closures. For example, during the H1N1 pandemic in Taiwan, teachers were on the forefront of alleviating the impact of school closures by contacting students to monitor their health, ensure they were practicing quarantine, and completing their schoolwork (Chen et al., 2011). During the COVID-19 pandemic lockdowns, educators took significant action to minimize learning disruptions by providing enough work for students to ensure they did not fall behind, and making themselves available for communication with students, while themselves isolating at home (Wang et al., 2020). Research examining teacher well-being during the lockdown in England found that teachers experienced a spike in work-related anxiety both when lockdowns were first announced and when they found out they would be returning to school (Allen et al., 2020), highlighting the impact of unpredictable changes due to the pandemic can have on teacher mental health. Nevertheless, Allen et al. (2020) found that while some aspects of overall teacher-well being decreased during lockdown, such as feeling useful or optimistic, other aspects improved, such as thinking clearly and having extra energy. Taken together, these studies suggest that the COVID-19 pandemic can impact teacher mental health in different ways, which in turn may influence their ability to effectively teach, especially from a distance.

The first COVID-19 pandemic-related shutdown is of particular importance to understand and examine as this was the first-time families and educators were faced with many new and unique challenges. In Ontario, a state of emergency was declared on March 17^th^, 2020, which came with the shutdowns of businesses, places of worship, and schools, and restrictions were placed on social gatherings (Rodrigues, 2020). Everyone was expected to stay home, except for accessing essentials, such as food, medicine, and healthcare. All schools across Ontario were closed during this time, remained closed for the rest of the school year (end of June 2020), and only reopened in September 2020 (CBC News, 2020). There were limited directives from the government regarding what learning should look like during this time compared to later on in the pandemic, resulting in a wide range of experiences of kindergarten learning across Ontario at this time (e.g., only posting assignments, multiple synchronous classes throughout the day, only 30-minute meetings, etc.). The state of emergency in Ontario remained in place until the end of July 2020 (Anandasagar, 2020) and there was a gradual loosening of restrictions during the summer months. This time in the pandemic resulted in increased challenges for families since many adults were faced with additional burdens of increased care-giving time, assisting in their children’s learning in a greater capacity, and helping older relatives navigate the restrictions.

### 1.3. Current Study

So far, studies of educator mental health have found somewhat conflicting results, with some researchers finding positive associations between the pandemic-related school closures and educator well-being, while others observing negative associations. Whereas previous research has begun to explore the association between caregiving and working from home during the COVID-19 pandemic, or examined teacher well-being in general, research has yet to explore the unique contribution of caregiving during the COVID-19 pandemic to the well-being of kindergarten educators. Due to their responsibility for teaching the youngest, least independent, school-age population, kindergarten educators faced particular challenges. Moreover, statistics show that they are themselves predominantly female and parents of school-age children (Government of Canada, 2011). The purpose of the current study was to therefore examine the association between caregiving responsibilities during the pandemic-related school closures with educators’ self-reported symptoms of depression and anxiety. We not only asked educators about their responsibilities of caring for children or an older adult, but due to the uniqueness of the pandemic-related school closures, we also asked educators if they were responsible for the learning of their own child(ren). This study was conducted using a web-based survey of kindergarten educators in Ontario, Canada, during the first provincial lockdown in the spring of 2020. We hypothesized that educators who reported being responsible for the care or learning of their own children, and those who looked after older adults would have higher levels of symptoms of depression and anxiety than those who did not have these additional responsibilities.

## 2. Methods

### 2.1. Study Design

Our study was a one-time, cross-sectional, population-based study of kindergarten educators working in publicly-funded schools in the province of Ontario, Canada.

### 2.2. Sample and Procedures

In Ontario, full-day kindergarten consists of a two-year, child-centered learning program for 4- and 5-year-old children where students attend school all day, five-days a week (*The Kindergarten Program*, 2016). The curriculum is focused on play-based learning, with the goal of promoting all aspects of their development, from physical health and well-being, to socioemotional and cognitive skills (Government of Ontario, 2019^b^). Since the introduction of full-day kindergarten in 2010, each class has an educator team, comprised of a teacher and an early childhood educator (ECE). Teachers have knowledge of the curriculum and are responsible for student learning (including assessments and evaluations) and reporting to parents. ECEs have knowledge of early childhood development and help promote children’s overall development by participating in the planning of age-appropriate activities (Government of Ontario, 2019^a^). Since kindergarten classes in Ontario, Canada are taught by two professionals, our study sample comprised both. Therefore, we refer to both teachers and early childhood educators jointly as “educators.” Invitations to participate in the study were sent out to kindergarten educators working at publicly-funded schools in the province of Ontario through their unions’ newsletters, as well as several other online publications reaching out to the target audience^1^. Given the kindergarten program’s emphasis on play-based learning, it was particularly difficult for kindergarten educators to effectively carry out the curriculum from a distance when schools were closed in the spring of 2020.

Participants completed a web-based survey entitled “Hidden Future Front Line: Educators’ perspective on the impact of the COVID-19 pandemic on kindergarten children (HiFLEC)” starting on May 26^th^ and ending on July 17^th^ 2020, thus at least 2 months after the beginning of school closures. Participants provided consent by clicking “yes” in the appropriate box located below the study information on the survey’s first page. Educators reported on a variety of topics regarding the COVID-19 pandemic including their own family situation, their experience with the transition to distance learning, and their own mental health. All methods and procedures were approved by the University Research Ethics Board.

### 2.3. Measures

#### 2.3.1. Demographics

Participants reported on demographic information, including their age, sex, years of experience as an educator, and household income.

#### 2.3.2. Family responsibilities

Educators reported on their current home life as it pertained to the COVID-19 pandemic. They were asked about family composition, and those with children under 18 years of age in their care were asked two additional questions regarding their children: Are you currently responsible for the 1) care, 2) learning of your own children? Response options were: Not primarily responsible/Not applicable; Yes, I share this responsibility with another adult; Yes, it is primarily my responsibility. All respondents were asked whether they were currently responsible for the care of older adults, with the following response options: Yes, living in the same household; Yes, living in another household; Yes, living in assisted care setting; No.

For childcare and learning, we compared three groups: 1) educators who reported that they either didn’t have young children or were not primarily responsible for their care or learning (this did not include educators who reported living on their own^2^); 2) educators who reported being primarily responsible for the care or learning of their own children and 3) educators who reported sharing these responsibilities with someone else. For adult care, we compared educators who reported caring for an older adult with educators that did not report this responsibility.

#### 2.3.3. Outcome measures: Educator mental health

Educator mental health was conceptualized as symptoms of depression and anxiety, measured with standardized short questionnaires.

##### Centre for Epidemiological Studies Depression Scale

(CESD-10; Radloff, 1977) This 10-item scale asks participants to rate on a scale of 0 to 3 how often over the past week they have felt symptoms associated with depression, where 0 = *rarely or none of the time* and 3 = *most or almost all of the time*. The CESD measures depressive symptomology in the general population. Higher scores on this scale indicate a higher level of depression. A score of 10 to 15 indicates moderate levels of depression and a score of 16 or higher is used for identifying those at risk for clinical depression (Lewinsohn et al., 1997). The sample-derived reliability for this scale was 0.89. Mean overall scores, as well as a cut-off score of 10 indicating at least moderate levels of depression (Lewinsohn et al., 1997), were used in the analyses.

##### General Anxiety Disorder Scale

(GAD; Spitzer et al., 2006) This is a 7-item scale that measures the severity of anxiety. Participants are asked: “Over the last two weeks, how often have you been bothered by the following” and responded on a scale from 0 = *not at all* to 3 = *nearly every day* with a sample-derived reliability of 0.82. Scores between 0 and 4 indicate minimal anxiety, 5 to 9 constitutes mild anxiety levels, 10 to 14 represents moderate levels of anxiety, and scores of 15 or higher indicate severe anxiety (Spitzer et al., 2006). Mean overall scores, as well as a cut-off score of 10 indicating at least moderate levels of anxiety (Spitzer et al., 2006), were used in the analyses.

### 2.4. Analytical Plan

Descriptive statistics (frequencies, percentages, means, and standard deviations) were used to examine the educators’ demographic characteristics (e.g., age, years of experience, family income) as well as how many educators reported having family responsibilities including caregiving and learning of children under the age of 18, and caring for an older adult. Next, we conducted three one-way multivariate analyses of covariance (MANCOVAs), one for each type of responsibilities, to examine whether there were significant differences on depression and anxiety scores between educators reporting having these responsibilities and those who did not. Finally, we conducted binary logistic regression analyses to establish the contribution of family responsibilities on the risk of meeting the criteria for at least moderate levels of anxiety and depression.

We controlled for respondent sex, educator years of experience, and family income in all analyses. Previous research suggests that there are significant differences in reported depression and anxiety in men and women, such that women tend to score higher on both outcomes (e.g., Faravelli et al., 2013). Further, previous research has suggested that higher levels of teacher effectiveness is associated with having more years of experience (e.g., Wolters & Daugherty, 2007), which is why years of experience variable was also included as a covariate in our analyses. Lastly, family income was important to include as a covariate given the association between lower socioeconomic status and higher levels of pandemic stress (Bambra et al., 2020; See Supplemental Table 2 for mean scores of depression and anxiety split by covariate variables). All preliminary and primary analyses were conducted using the statistical software IBM SPSS version 25 (IBM Corp., 2017).

## 3. Results

### 3.1. Demographic Characteristics of Educators

A total of 2,189 educators had valid responses on the depression items, and 2,175 on anxiety (2,173 had valid scores on both). To be included in further analyses, they also had to have valid responses on the sex, work experience, and family care variables. This resulted in a final analytical sample of 1,790. The mean scores and prevalence rates of the mental health measures in the final sample were not markedly different from the full sample (See Supplemental Table 1).

Of the educators, 73.6% were kindergarten teachers and 26.4% were early childhood educators. Educators who responded represented 74 of the 75 school districts/authorities across Ontario. Experience as either a kindergarten teacher or early childhood educator ranged from less than 5 years to 21 years or more. See Table 1 for breakdown of sample characteristics.

**Table 1.**
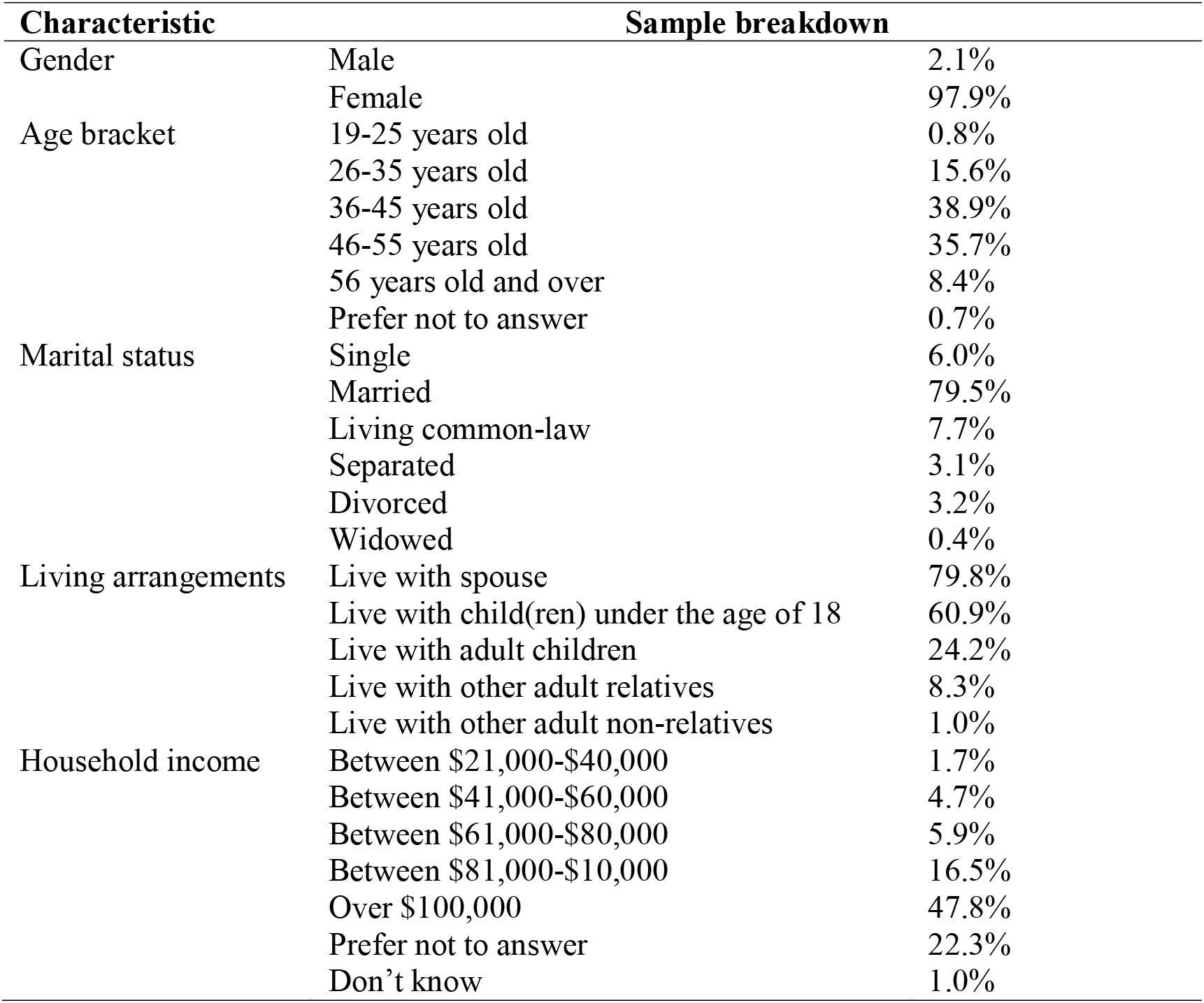
*Sociodemographic characteristics of the study sample* (N = 1,790)

### 3.2. Family Responsibilities

Educators were asked about their family responsibilities during the COVID-19 pandemic. Figure 1 demonstrates the proportion of educators in each group of interest. Figure 1A displays percentages of educators who reported they did not have the responsibility, shared the responsibility, or were primarily responsible for caregiving or learning of children under the age of 18 years (39.9%, 37.4%, 22.7%; 42.0%, 19.6%, 38.4%, respectively); Figure 1B displays percentages of educators who reported either not having or having the responsibility of caring for another adult (66% and 34% respectively).

**Figure 1.**
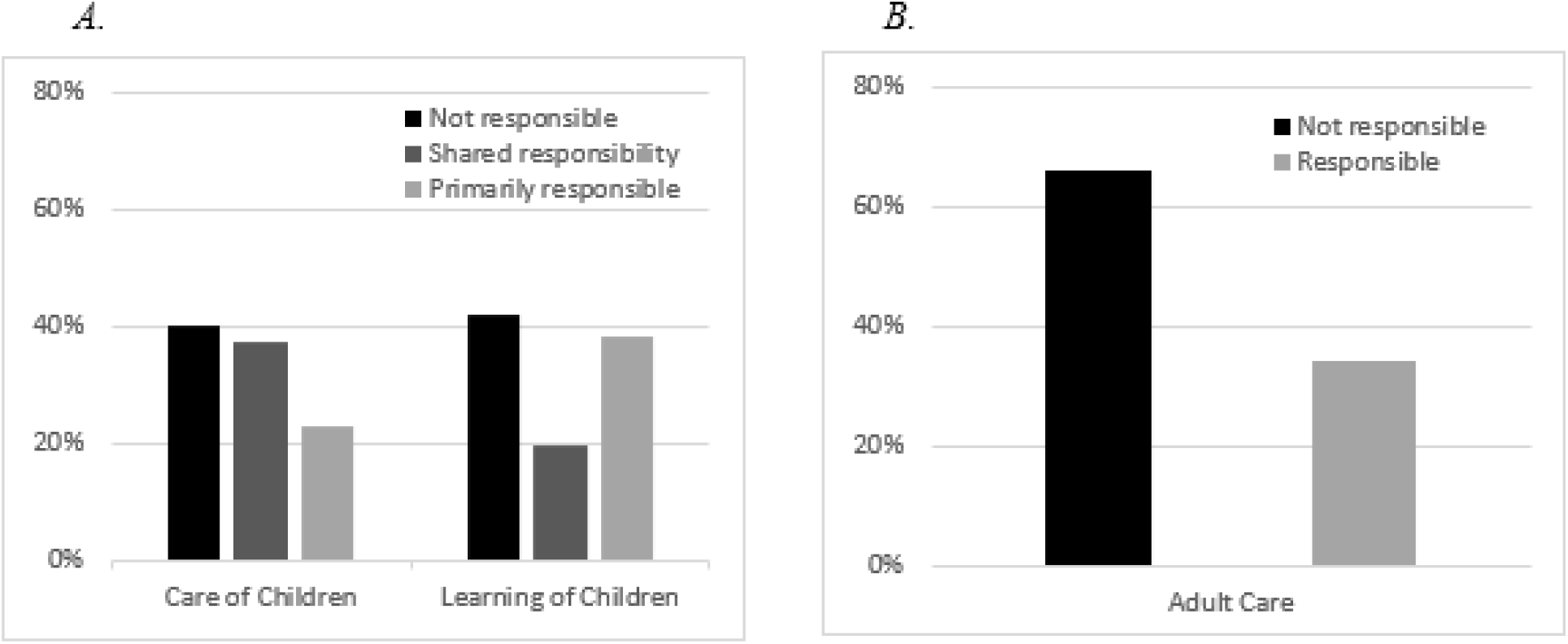
Percentages of educators by type of responsibility for the care or learning of their own children or care of older adults

### 3.3. Mental Health Status

Study respondents had a mean depression score of 11.45 (*SD* = 6.74) and a mean anxiety score of 7.04 (*SD* = 5.60). Of the educators in our sample, 50.4% scored above the moderate level cut-off on depression, 24.5% scored above the moderate level cut-off for anxiety, and 22.7% had scores above the moderate level cut-off for both depression and anxiety. Proportions of educators who scored above the moderate level cut-offs for depression and anxiety in each family responsibility category are shown in Table 2. Overall, there were higher proportions of educators with depression and anxiety scores above the moderate level cut-offs among those who were primarily responsible for childcare or learning, and those who cared for older adults than among those who did not.

**Table 2.** Proportions of educators who scored above the moderate level cut-off for anxious and depressive symptoms by family responsibility

|  | % above cut-off |  |
| --- | --- | --- |
|  | Depression | Anxiety |
| Childcare |  |  |
| No | 47.6% | 23.6% |
| Shared | 47.1% | 23.3% |
| Primarily | 60.8% | 27.8% |
| Child Learning |  |  |
| No | 48.2% | 23.4% |
| Shared | 43.7% | 23.7% |
| Primarily | 56.0% | 25.9% |
| Adult Care |  |  |
| No | 48.2% | 21.6% |
| Yes | 54.6% | 30.1% |

### 3.4. Association Between Educators’ Mental Health and Family Responsibilities

We conducted three MANCOVA analyses to investigate the impact of having added family responsibilities (childcare, child learning, and adult care) on educator mental health. Our intent was to control for respondent sex, years of experience, and income in each analysis, however, 23.3% of educators had missing data on the household income variable (responded “prefer not to say” or “don’t know”). We conducted the analyses both with and without the income variable and found that patterns of main results were consistent (results including income as a covariate are presented in Supplemental Tables 3-5). Therefore, here we present results controlling only for respondent sex and years of experience.

#### 3.4.1. Childcare Responsibilities

Results of a MANCOVA examining the association between childcare responsibilities and symptoms of depression and anxiety showed significant differences between the three groups (Pillai’s trace = .014, F (4, 3570) = 6.23, *p* < .001). Bonferroni adjustment for multiple comparisons revealed that educators who reported being primarily responsible for the caregiving of their own children had significantly higher scores on depression than those reporting they shared responsibilities or did not have these responsibilities. There were no significant differences between the group who shared responsibilities and those who did not have caregiving responsibilities (Table 3). There were no significant differences among the three groups on symptoms of anxiety.

**Table 3.** MANCOVA results examining differences in depression and anxiety scores among educators who reported primary, shared, and no responsibilities for childcare

| | Mean | S.E. | F | P | Partial $\eta^2$ |
| --- | --- | --- | --- | --- | --- |
| Depression |  |  |  |  |  |
| Responsible for childcare |  |  |  |  |  |
| No/not applicable | 11.31 | .25 | F (2, 487.62) = 10.85 | <.001 | .012 |
| Shared responsibility | 10.81 | .26 |  |  |  |
| Primarily responsible | 12.76 | .33 |  |  |  |
| Anxiety |  |  |  |  |  |
| Responsible for childcare |  |  |  |  |  |
| No/not applicable | 6.89 | .21 | F (2, 93.78) = 3.01 | .050 | .003 |
| Shared responsibility | 6.85 | .25 |  |  |  |
| Primarily responsible | 7.64 | .28 |  |  |  |

*Note.* MANCOVA controlled for sex and years of experience. Comparisons based on estimated marginal means. Adjusted for Bonferroni multiple comparisons.

#### 3.4.2. Learning Responsibilities

The second MANCOVA compared educators’ symptoms of depression and anxiety in relation to being responsible for the learning of their own children. Results of this analysis revealed that there were significant group differences (Pillai’s trace = .006, F (4, 3566) = 3.53, *p* = .022). Similar to childcare responsibilities, there were only statistically significant differences between the groups on symptoms of depression (Table 4). Comparisons based on estimated marginal means and using a Bonferroni adjustment for multiple comparisons revealed that educators who reported they were primarily responsible for their own child’s learning had significantly higher depression scores than those who reported that they shared this responsibility, but did not differ from educators who did not have this responsibility. Once again, there were no significant differences between the three groups on symptoms of anxiety.

**Table 4.** MANCOVA results examining differences in scores of depression and anxiety among educators who reported primary, shared, and no responsibilities for child learning

| | Mean | S.E. | F | P | Partial $\eta^2$ |
| --- | --- | --- | --- | --- | --- |
| Depression |  |  |  |  |  |
| Responsible for child learning |  |  |  |  |  |
| No/not applicable | 11.34 | .25 | F (2, 230.44) = 5.09 | .006 | .006 |
| Shared responsibility | 10.61 | .36 |  |  |  |
| Primarily responsible | 12.00 | .26 |  |  |  |
| Anxiety |  |  |  |  |  |
| Responsible for child learning |  |  |  |  |  |
| No/not applicable | 6.87 | .20 | F (2, 74.68) = 2.39 | .092 | .003 |
| Shared responsibility | 6.70 | .30 |  |  |  |
| Primarily responsible | 7.40 | .21 |  |  |  |

*Note.* MANCOVA controlled for sex and years of experience. Comparisons based on estimated marginal means. Adjusted for Bonferroni multiple comparisons.

#### 3.4.3. Adult Care Responsibilities

Results of the third MANCOVA examining the association between caring for an older adult and symptoms of depression and anxiety indicated that there were significant differences between educators who reported that they were responsible for the care of older adults and those who were not (Pillai’s trace = .013, F (2, 1925) = 13.05, *p* < .001). As can be seen in Table 5, there were significant differences between these two groups on both symptoms of depression and anxiety. Educators who reported caring for an older adult had higher levels of depression and anxiety compared to those who did not.

**Table 5.** MANCOVA results examining differences in scores of depression and anxiety between educators who reported adult care responsibilities and those who did not

| | Mean | S.E. | F | P | Partial $\eta^2$ |
| --- | --- | --- | --- | --- | --- |
| Depression |  |  |  |  |  |
| Responsible for older adult |  |  |  |  |  |
| No | 11.09 | .19 | F (1, 712.90) = 15.87 | <.001 | .008 |
| Yes | 12.41 | .27 |  |  |  |
| Anxiety |  |  |  |  |  |
| Responsible for older adult |  |  |  |  |  |
| No | 6.56 | .16 | F (1, 808.57) = 26.01 | <.001 | .013 |
| Yes | 7.96 | .22 |  |  |  |

*Note.* MANCOVA controlled for sex and years of experience. Comparisons based on estimated marginal means. Adjusted for Bonferroni multiple comparisons.

### 3.5. Association Between Any Family Responsibilities and Educator Mental Health

In order to determine the comparative strength of association between any of the family responsibilities and scoring above the moderate level cut-off for depression and anxiety, we conducted two binary logistic regressions (Table 6). Adjusted analyses that included respondent sex, years of experience, and income as covariates showed main effects consistent with the unadjusted models. For the current analyses, we split years of experience into two groups by the median range of years: up to 15 and 16 or more. Overall, 46.9% had 0-15 years of experience, while 53.1% had 16 or more years of experience in their respective profession.

**Table 6.** Results of unadjusted and adjusted binary logistic regressions examining the association between family responsibilities and symptoms of depression and anxiety

|  | Unadjusted OR (95% CI) |  | Adjusted OR (95% CI) |  |
| --- | --- | --- | --- | --- |
| Depression |  |  |  |  |
| Childcare (Primary) | 2.21* | (1.16 – 4.18) | 2.14* | (1.13 – 4.07) |
| Childcare (Shared) | 1.43 | (.78 – 2.64) | 1.41 | (.76 – 2.59) |
| Child learning (Primary) | .79 | (.43 – 1.45) | .80 | (.43 – 1.48) |
| Child learning (Shared) | .60 | (.32 – 1.12) | .62 | (.33 – 1.17) |
| Older adult care | 1.33* | (1.09 – 1.62) | 1.34* | (1.12 – 1.68) |
| Sex |  |  | .52 | (.26 – 1.03) |
| Experience |  |  | .86 | (.71 – 1.04) |
| Anxiety |  |  |  |  |
| Childcare (Primary) | 1.06 | (.49 – 2.30) | 1.02 | (.47 – 2.20) |
| Childcare (Shared) | .83 | (.40 – 1.75) | .82 | (.39 – 1.74) |
| Child learning (Primary) | 1.21 | (.58 – 2.55) | 1.26 | (.60 – 2.66) |
| Child learning (Shared) | 1.24 | (.58 – 2.65) | 1.27 | (.59 – 2.74) |
| Older adult care | 1.57* | (1.26 – 1.97) | 1.67* | (1.33 – 2.10) |
| Sex |  |  | .92 | (.43 – 1.97) |
| Experience |  |  | .74* | (.59 – .93) |

*Note.* Adjusted analyses controlled for sex and years of experience. Reference groups for all care/learning variables was the “no/not responsible” group. Reference group for sex was female. Reference group for Experience was 0-15 years.

As with previous analyses, only sex and years of experience were entered as covariates to preserve sample size (see Table 6). Results remained consistent when income variables were included (see Table S2). Educators had 2.14 times greater odds of scoring above the cut-off on depressive symptoms if they had the responsibility of caring for children under the age of 18 years and 1.34 times greater odds if they were responsible for caring for an older adult. Educators were also 1.67 times more likely to score above the cut-off for anxiety if they had the responsibility of caring for an older adult.

## 4. Discussion

Our study examined the mental health of kindergarten educators’ in Ontario, Canada, during the COVID-19-related school closures in the spring of 2020. The goal of the current study was to examine the association between kindergarten educators’ responsibility for the care or learning of their own children and the care of older adults and their self-reported symptoms of depression and anxiety. Overall, our results supported our hypotheses, as educators who reported caring responsibilities had poorer mental health scores, however, not all these differences reached statistical significance. Educators who reported being primarily responsible for the care and learning of their own children had statistically significantly higher levels of depression, while those who were responsible for the care of older adults had statistically significantly higher levels of both anxiety and depression than those who did not.

When the first wave of public health responses to the COVID-19 pandemic was implemented in the spring of 2020 in Ontario, it included immediate school closures, forcing educators to quickly adjust to teaching online, and for many of them (about 60% of our sample) at the same time also helping their own children adjust to learning from home. Even though current evidence suggests that teacher mental health has been negatively impacted by the added stress of distance teaching during school closures (e.g., Alves et al., 2020), no research to date investigated the impact of family responsibilities on educator mental health in these circumstances. We found significant differences in levels of depression between the groups of educators who reported primary, shared, and no childcare responsibility. For both depression and anxiety, mean scores were highest for the group of educators who reported that they were primarily responsible for the care of their own children under the age of 18 years, compared to both the group that reported that they shared or did not have this responsibility, however these differences were not significant for anxiety. These results align with existing research on family responsibilities. During the COVID-19 pandemic, parents in China who had children in primary or secondary school had higher rates of depression and anxiety compared to parents who had college-aged children (Wu et al., 2020). Research conducted between May and November 2020 in Canada found that adults with children had higher levels of anxiety than Canadians who did not have children (CAMH, 2020), even though, similar to our study, the results did not consistently reach statistical significance. Our findings also support the notion that working parents may be struggling with balancing working from home while also having their children at home, which may be contributing to poorer mental health outcomes (e.g., Patrick et al., 2020).

Our results also help clarify the previous mixed results regarding association between childcare and teacher mental health during the COVID-19 pandemic (e.g., Allen et al., 2020). Specifically, though not significantly different, the group of educators that reported that they did not have the responsibility for the care or learning of children consistently had poorer mental health compared to those who reported sharing this responsibility. While these results were initially unexpected, they support research by Lades and colleagues (2020) suggesting that there may be benefits during the pandemic to the well-being of adults who have children to care for. It appears that having children to focus on and care for may help mitigate feelings of depression and anxiety. However, out data suggest that this only appears to be the case if that responsibility is shared with another adult. The fact remains, that educators who reported being primarily responsible for the care or learning of their own children had the highest levels of both depression and anxiety. Our study is the first to our knowledge that has examined this difference between primary and shared child responsibilities.

During the school closures in Ontario from March-June 2020, many educators also had to support their own child with their online distance learning. This challenge has been a highly unique aspect of the pandemic-related school closures, and thus our exploration of its impact on educators is a unique contribution of the current study. While caregiving responsibilities of young children is a fact of life for parents, the pandemic-related shutdowns of businesses and schools meant that employed parents had to fulfill all their various roles at the same time. They had to care for their school-aged children and ensure that they were engaging in online learning while also trying to work themselves. Our data revealed that 38.4% of educators were primarily responsible for the learning of their own child(ren), compared to 22.7% of educators who reported being primarily responsible for the care of their children. This suggests that even if educators have a partner that shares the caregiving responsibilities, a greater percentage of educators are still primarily responsible for their own child(ren)’s learning. To our knowledge, no other study explored the differential burden of these two responsibilities in any group of parents working from home during the COVID-19 pandemic. Even though educators who were primarily responsible for their own children’s learning tended to report higher levels of anxiety than those who shared the responsibility or had none, these differences were not statistically significant. We also found that the burden of responsibility of caring for older adults carried a risk of increased levels of depression and anxiety. This is consistent with Kent et al.’s study (2020) who found that individuals responsible for caring for elderly relatives reported increased stress due to the COVID-19 pandemic, which in turn resulted in poorer mental health outcomes.

When we examined the contribution of both types of responsibilities to educator mental health, we found that caring for both a child and an older adult was significantly associated with scoring above the cut-off for moderate levels of depression. However, having the responsibility of caring for a child had a higher odds ratio (2.14) than caring for an older adult (1.34). In contrast, when considered together, only caregiving for an older adult significantly increased an educator’s odds of reporting at least moderate symptoms of anxiety. These results align with previous research, even though these two types of responsibilities had not been previously considered together. In a sample of parents of children under 18 years in the United States, Brown et al. (2020) reported that elevated stress due to COVID-19 pandemic was associated with higher anxiety and depression. The burden of caregiving for older family members prompted concerns among professionals about the mental health of and required supports for caregivers (Kent et al., 2020). The increased anxiety seen in caregivers of older adults likely stemmed from the early knowledge of the consequences of the COVID-19 virus infection, which emphasized a high health risk for older adults, but little risk for children (Davies et al., 2020). Research has also suggested that fear of contracting the virus is associated with poorer psychological well-being in older adults (Whitehead & Torossian, 2021), a fear that is likely shared by their caregivers. Therefore, educators who were responsible for the care of an older adult may have been more anxious because of needing to navigate how to care for family members while balancing isolation measures (Kent et al., 2020).

Given the increasing demands on educators in both their professional and home life during school closures, it is important to consider their own well-being. For example, the school closures saw educators facing challenges associated with isolation measures, such as post-traumatic stress symptoms, experiencing fears of infection, frustration with uncertainty, and decreased social and physical contact with others (Brooks et al., 2020). This is in addition to distance learning challenges, as well as work-home life balance, all of which can impact mental health (Strauss, 2020). Our results suggest that this might be particularly salient for educators who are responsible for the care and learning support of their own children, or for the care of an older adult in their family, while also having to teach the students in their class from home. Literature to date on the COVID-19 pandemic has suggested that working parents are faced with added stress of navigating their caregiving responsibilities and their professional obligations, however much of this research has been focused on front-line workers (e.g., Varner, 2020). In our sample, there was only about a 23% overlap between moderate-or-higher symptomatology between depression and anxiety, whereas the comorbidity of those two disorders is generally being reported at much higher rates, between 60-70% (e.g., Groen et al., 2020). We interpreted our low rates of overlap to indicate that, in our sample, these two mental health issues were fairly distinct, and therefore did not examine a combined outcome of depression and anxiety.

It should be noted that the kindergarten educators in our sample were predominantly female. Since teaching is a profession dominated by women, particularly in elementary grades,^3^ our findings have particular relevance to working women. To date, research has pointed to the pandemic having a larger impact on working women compared to working men, especially those with children. Research focused on educator well-being during the pandemic found that female teachers experienced higher levels of stress due to school closures, compared to their male counterparts, which they attributed to higher workload and increased domestic responsibilities (Allen et al., 2020; Klapproth et al., 2020). Recent research has also suggested that higher demands in teaching due to the increased technology use have been particularly challenging for teachers who are mothers, due to having less free time and having blurred boundaries between their roles at home and at work (Weisberger et al., 2021). Further, women are reported to be taking on the majority of the extra child and home care due to the pandemic shutdowns (e.g., Power, 2020; Quian & Fuller, 2020; Yamamura & Tsustsui, 2021). Power (2020) suggests that although women were already taking on most of the unpaid work prior to the pandemic, the proportion has dramatically increased since March 2020 when the pandemic began, which may have long-term negative impacts on women and families for several years to come. For instance, the employment gender gap has significantly widened as a result of the COVID-19 pandemic among parents of elementary school-aged children, and women have reported lower productivity and job satisfaction compared to men (e.g., Feng & Savani, 2020; Quian & Fuller, 2020; Yamamura & Tsustui, 2021).

### 4.1. Strengths, Limitations and Future Directions

To our knowledge, our study is the first to investigate the associations between family responsibilities and mental health outcomes of a large sample of kindergarten educators during the COVID-19 pandemic. Our study’s major strength is that it demonstrates the burden of multiple responsibilities and its association with poor mental health among educators responsible for teaching children in the youngest elementary school grades.

Nevertheless, our study has a number of limitations that should be mentioned. Whereas a strength of our study is that our respondents came from all but one Ontario school division/authority, it was not possible for us to assess the representativeness of this sample. Because our study only included kindergarten educators, it is likely that these findings might differ for teachers of children in older grades. Future research should examine similar associations in teachers of all grade levels. Moreover, our study was cross-sectional and therefore we could not establish a causal relationship between family responsibility and symptoms of depression and anxiety. We were also unable to examine whether any pre-existing mental health conditions may have contributed to our results. Our study was conducted at a specific period in time and our results can only be interpreted in the context of the spring of 2020, during the first round of the COVID-19-related school closures in Ontario, Canada. As the landscape of the COVID-19 pandemic continues to change with respect to schools, learning (e.g., Sokal et al. 2020), and the adults’ and children’s susceptibility to the illness, research should continue to examine educator mental health, particularly from the perspective of balancing family responsibilities and remote teaching.

### 4.2. Conclusion

The results of the current study are an important addition to the existing literature on distance teaching during the COVID-19 pandemic-related school closures, and on the burden of family responsibilities on professionals working from home, exemplified by the experience of kindergarten educators. Overall, our study suggests that having family responsibilities of caregiving or learning of children under the age of 18 years or caring for older adults during the COVID-19 pandemic, especially when these responsibilities were not shared, were negatively associated with educator mental health. This is especially important considering kindergarten educators are predominantly women, who tend to be taking on most of the extra family care responsibilities (e.g., Power, 2020). In light of the ongoing COVID-19 pandemic, it is important to systematically investigate the context of the associated school closures, and the impact that this had on educators, caregivers, and their families. While most of such research focuses on the impact on children, the current work provides insight into the first pandemic-related shutdown, the first time that educators were faced with the challenges of teaching from home, with increased responsibilities due to the pandemic. These findings can also help inform future research exploring pandemic-related shutdowns. Since this study was conducted, the province of Ontario has been through two additional lockdowns that have also impacted families and may also have been associated with changes in their mental health and well-being. Pre-pandemic research has suggested that organizations should develop programs that promote a work-life balance for teachers who are also parents (Palmer et al., 2012). Our results indicate that this may be even more important during the COVID-19 pandemic. Altogether, these results emphasize the need for mental health supports not only for educators, but also for parents of school-aged children who work from home during the school closures.

## Data Availability

Data available upon reasonable request and partner consent

---

**Table S1.** Comparison of depression and anxiety scores between the total study sample with valid scores and the final analytical sample

| **Score** | **Whole Sample with valid depression or anxiety score** | **Study Sample** |
| --- | --- | --- |
| Mean depression score (*SD*) | 11.49 (6.75) | 11.45 (6.74) |
| Mean anxiety score (*SD*) | 6.96 (5.66) | 7.04 (5.60) |
| % above cut-off on depression | 50.8% | 50.4% |
| % above cut-off on anxiety | 24.4% | 24.5% |

**Table S2.**
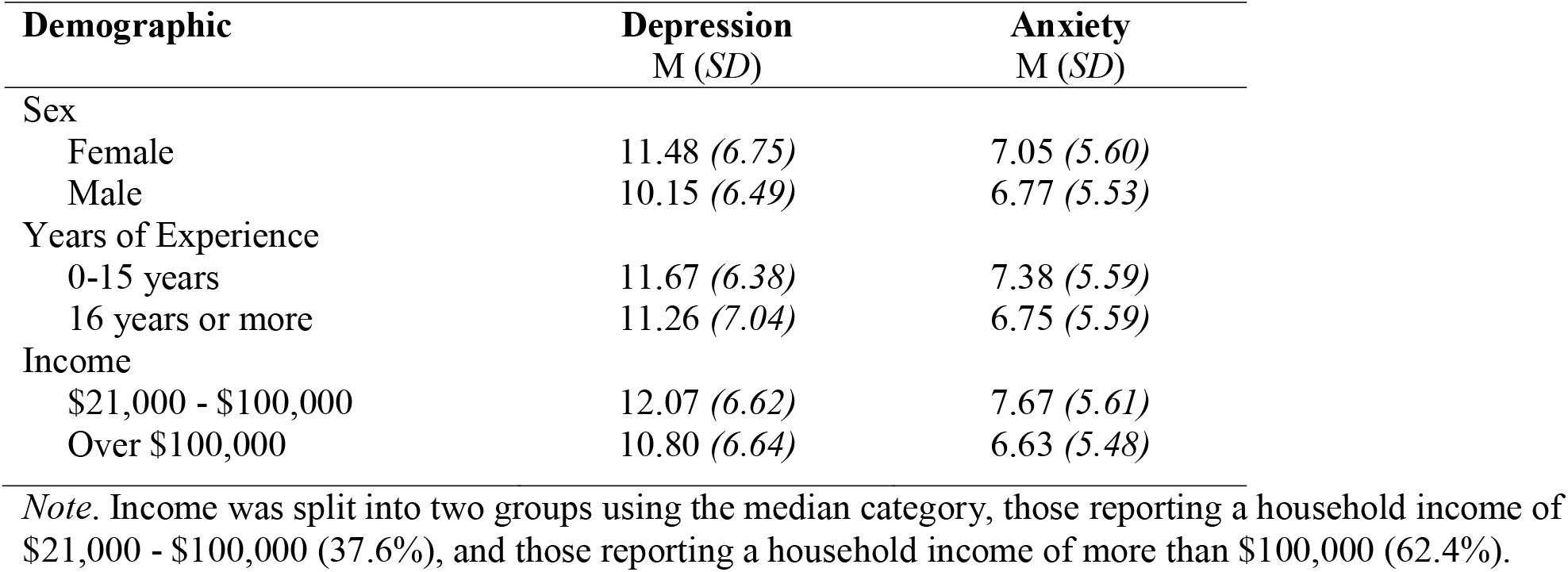
Mean depression and anxiety scores by educators’ demographic covariates

| **Demographic** | **Depression**<br>**M (*SD*)** | **Anxiety**<br>**M (*SD*)** |
| --- | --- | --- |
| Sex |  |  |
| Female | 11.48 (6.75) | 7.05 (5.60) |
| Male | 10.15 (6.49) | 6.77 (5.53) |
| Years of Experience |  |  |
| 0-15 years | 11.67 (6.38) | 7.38 (5.59) |
| 16 years or more | 11.26 (7.04) | 6.75 (5.59) |
| Income |  |  |
| \$21,000 - \$100,000 | 12.07 (6.62) | 7.67 (5.61) |
| Over \$100,000 | 10.80 (6.64) | 6.63 (5.48) |

*Note.* Income was split into two groups using the median category, those reporting a household income of \$21,000 - \$100,000 (37.6%), and those reporting a household income of more than \$100,000 (62.4%).

**Table S3.** MANCOVA results examining differences in depression and anxiety scores among educators who reported primary, shared, and no responsibilities for childcare

| | Mean | S.E. | F | P | Partial $\eta^2$ |
| --- | --- | --- | --- | --- | --- |
| Depression |  |  |  |  |  |
| Responsible for childcare |  |  |  |  |  |
| No/not applicable | 11.19 | .29 | F (2, 370.02) = 8.51 | <.001 | .012 |
| Shared responsibility | 10.58 | .30 |  |  |  |
| Primarily responsible | 12.53 | .37 |  |  |  |
| Anxiety |  |  |  |  |  |
| Responsible for childcare |  |  |  |  |  |
| No/not applicable | 7.00 | .24 | F (2, 30.84) = 1.01 | .364 | .001 |
| Shared responsibility | 6.82 | .25 |  |  |  |
| Primarily responsible | 7.38 | .31 |  |  |  |

*Note.* MANCOVA controlled for sex and years of experience. Comparisons based on estimated marginal means. Adjusted for Bonferroni multiple comparisons.

**Table S4.** MANCOVA results examining differences in scores of depression and anxiety among educators who reported primary, shared, and no responsibilities for child learning

| | Mean | S.E. | F | P | Partial $\eta^2$ |
| --- | --- | --- | --- | --- | --- |
| Depression |  |  |  |  |  |
| Responsible for child learning |  |  |  |  |  |
| No/not applicable | 11.22 | .28 | F (2, 296.93) = 6.81 | .001 | .010 |
| Shared responsibility | 10.10 | .41 |  |  |  |
| Primarily responsible | 11.94 | .29 |  |  |  |
| Anxiety |  |  |  |  |  |
| Responsible for child learning |  |  |  |  |  |
| No/not applicable | 6.99 | .23 | F (2, 61.21) = 2.01 | .135 | .003 |
| Shared responsibility | 6.49 | .34 |  |  |  |
| Primarily responsible | 7.33 | .24 |  |  |  |

*Note.* MANCOVA controlled for sex and years of experience. Comparisons based on estimated marginal means. Adjusted for Bonferroni multiple comparisons.

**Table S5.** MANCOVA results examining differences in scores of depression and anxiety between educators who reported adult care responsibilities and those who did not

| | Mean | S.E. | F | P | Partial $\eta^2$ |
| --- | --- | --- | --- | --- | --- |
| Depression |  |  |  |  |  |
| Responsible for older adult |  |  |  |  |  |
| No | 10.98 | .21 | F (1, 448.09) = 10.31 | .001 | .007 |
| Yes | 12.17 | .30 |  |  |  |
| Anxiety |  |  |  |  |  |
| Responsible for older adult |  |  |  |  |  |
| No | 6.48 | .18 | F (1, 810.69) = 26.83 | <.001 | .018 |
| Yes | 8.09 | .25 |  |  |  |

*Note.* MANCOVA controlled for sex and years of experience. Comparisons based on estimated marginal means. Adjusted for Bonferroni multiple comparisons.

**Table S6.** Results of unadjusted and adjusted binary logistic regression of the association between family responsibilities and moderate levels of depression and anxiety, including income as a covariate

|  | Unadjusted OR (95% CI) |  | Adjusted OR (95% CI) |  |
| --- | --- | --- | --- | --- |
| Depression |  |  |  |  |
| Childcare (Primary) | 2.28* | (1.12 – 4.67) | 2.24* | (1.09 – 4.59) |
| Childcare (Shared) | 1.50 | (.75 – 2.99) | 1.57 | (.78 – 3.14) |
| Child learning (Primary) | .76 | (.38 – 1.50) | .76 | (.38 – 1.52) |
| Child learning (Shared) | .51 | (.25 – 1.03) | .52 | (.25 – 1.06) |
| Older adult care | 1.26* | (1.01 – 1.58) | 1.31* | (1.04 – 1.66) |
| Sex |  |  | .70 | (.33 – 1.46) |
| Experience |  |  | .79* | (.63 – .98) |
| Income |  |  | .74* | (.59 – .92) |
| Anxiety |  |  |  |  |
| Childcare (Primary) | 1.03 | (.44 – 2.42) | 1.01 | (.43 – 2.37) |
| Childcare (Shared) | .87 | (.38 – 1.99) | .92 | (.40 – 2.13) |
| Child learning (Primary) | 1.05 | (.46 – 2.39) | 1.07 | (.47 – 2.44) |
| Child learning (Shared) | .96 | (.41 – 2.26) | .96 | (.40 – 2.27) |
| Older adult care | 1.84* | (1.43 – 2.38) | 1.98* | (1.52 – 2.58) |
| Sex |  |  | 1.30 | (.58 – 2.89) |
| Experience |  |  | .67* | (.53 – .89) |
| Income |  |  | .74* | (.57 – .95) |

*Note.* Adjusted analyses controlled for sex and years of experience. Reference groups for all care/learning variables was the “no/not responsible” group. Reference group for sex was female. Reference group for Experience was 0-15 years.

## Footnotes

1 About 92% of school aged children are enrolled in public schools across Canada (Government of Canada, 2018), and so private schools were not included in the current sample.)

2 For the purposes of the current study, we did not include educators who reported that they lived alone to focus our analyses on the potential implications of family responsibilities. Previous literature has suggested that individuals who were living alone during the pandemic experienced a heightened risk of loneliness and mental health challenges due to the COVID-19 pandemic (e.g., Bu et al., 2020).

3 Statistics Canada reports that as of 2011, 84% of all elementary and kindergarten teachers and 97% of all early childhood educators and assistants in Canada were women (Government of Canada, 2011).

## References

Allen, R., Jerrim, J., & Simms, S. (2020). How did the early stages of the COVID-19 pandemic affect teacher wellbeing?. Centre for Education Policy and Equalising Opportunities (CEPEO) Working Paper, (20-15), 20–15.

Altieri, M., & Santangelo, G. (2021). The psychological impact of COVID-19 pandemic and lockdown on caregivers of people with dementia. The American Journal of Geriatric Psychiatry, 29(1), 27–34.

Anandasagar, T. (2020). Bill 195: Ontario ends declared emergency, continues some emergency orders. Gowlingwlg.com. Gowling WLG International Limited. July 28, 2020. Retrieved September 19, 2021.

Anxiety patterns in Canadians mirror progression of pandemic. (2020, December 15). Retrieved February 26, 2021, from https://www.camh.ca/en/camh-news-and-stories/anxiety-patterns-in-canadians-mirror-progression-of-pandemic

Bambra, C., Riordan, R., Ford, J., & Matthews, F. (2020). The COVID-19 pandemic and health inequalities. J Epidemiology Community Health, 74(11), 964–968.

Baykal, S., Karakurt, M. N., Çakır, M., & Karabekiroğlu, K. (2019). An examination of the relations between symptom distributions in children diagnosed with autism and caregiver burden, anxiety and depression levels. Community Mental Health Journal, 55(2), 311–317.

Brooks, S. K., Webster, R. K., Smith, L. E., Woodland, L., Wessely, S., Greenberg, N., & Rubin, G. J. (2020). The psychological impact of quarantine and how to reduce it: rapid review of the evidence. The Lancet, 395(10227), 912–920.

Brown, S. M., Doom, J. R., Lechuga-Peña, S., Watamura, S. E., & Koppels, T. (2020). Stress and parenting during the global COVID-19 pandemic. Child Abuse & Neglect, 110, (104699). https://doi.org/10.1016/j.chiabu.2020.104699

Bu, F., Steptoe, A., & Fancourt, D. (2020). Who is lonely in lockdown? Cross-cohort analyses of predictors of loneliness before and during the COVID-19 pandemic. Public Health, 186, 31–34.

Burić, I., Slišković, A., & Penezić, Z. (2019). Understanding teacher well-being: a cross-lagged analysis of burnout, negative student-related emotions, psychopathological symptoms, and resilience. Educational Psychology, 39 (9), 1136–1155.

CBC News. Ontario shuts schools until September because of COVID-19 pandemic”. CBC News. May 19, 2020. Archived from the original on May 19, 2020. Retrieved September 19, 2021.

Chang, H. Y., Chiou, C. J., & Chen, N. S. (2010). Impact of mental health and caregiver burden on family caregivers’ physical health. Archives of gerontology and geriatrics, 50(3), 267–271.

Chen, W.C., Huang, A.S., Chuang, J.H., Chiu C.C., Kuo, H.S. (2011). Social and economic impact of school closure resulting from pandemic influenza A/H1N1. Journal of Infection, 62(3), 200–203.

Davies, N. G., Klepac, P., Liu, Y., Prem, K., Jit, M., & Eggo, R. M. (2020). Age-dependent effects in the transmission and control of COVID-19 epidemics. Nature Medicine, 26(8), 1205–1211.

Government of Canada. (2017, September 15). Back to school… by the numbers. Government of Canada, Statistics Canada. https://www.statcan.gc.ca/eng/dai/smr08/2014/smr08_190_2014#a5.

Government of Canada, S. C. (2018). The Daily—Elementary–Secondary Education Survey for Canada, the provinces and territories, 2016/2017. https://www150.statcan.gc.ca/n1/daily-quotidien/181102/dq181102c-eng.htm

^a^Government of Ontario (2019). Full-day kindergarten. Retrieved April 15, 2021, from http://www.edu.gov.on.ca/kindergarten/whoisworkingintheclassroom.html

^b^Government of Ontario (2019). The Ontario Curriculum: Elementary. Retrieved August 19, 2021, from http://www.edu.gov.on.ca/eng/curriculum/elementary/kindergarten.html

Groen, R. N., Ryan, O., Wigman, J. T., Riese, H., Penninx, B. W., Giltay, E. J., … & Hartman, C. A. (2020). Comorbidity between depression and anxiety: Assessing the role of bridge mental states in dynamic psychological networks. BMC Medicine, 18(1), 1–17.

Faravelli, C., Scarpato, M. A., Castellini, G., & Sauro, C. L. (2013). Gender differences in depression and anxiety: the role of age. Psychiatry Research, 210(3), 1301–1303.

Fauzi, I., & Khusuma, I. H. S. (2020). Teachers’ elementary school in online learning of COVID-19 pandemic conditions. Jurnal Iqra’: Kajian Ilmu Pendidikan, 5(1), 58–70.

Feng, Z., & Savani, K. (2020). Covid-19 created a gender gap in perceived work productivity and job satisfaction: implications for dual-career parents working from home. Gender in Management, 35, 719–736.

Jeffords, S. (2020, April 06). Ontario students begin e-learning as Covid-19 Closures continue. Retrieved March 04, 2021, from https://toronto.ctvnews.ca/ontario-students-begin-e-learning-as-covid-19-closures-continue-1.4883925

Hastings, R. P., & Bham, M. S. (2003). The relationship between student behaviour patterns and teacher burnout. School Psychology International, 24(1), 115–127.

Kent, E. E., Ornstein, K. A., & Dionne-Odom, J. N. (2020). The family caregiving crisis meets an actual pandemic. Journal of pain and symptom management, 60 (1), 66–69.

Klapproth, F., Federkeil, L., Heinschke, F., & Jungmann, T. (2020). Teachers’ experiences of stress and their coping strategies during COVID-19 induced distance teaching. Journal of Pedagogical Research, 4(4), 444–452.

Košir, K., Dugonik, Š., Huskić, A., Gračner, J., Kokol, Z., & Krajnc, ž. (2020). Predictors of perceived teachers’ and school counsellors’ work stress in the transition period of online education in schools during the COVID-19 pandemic. Educational Studies, 1–5. https://doi.org/10.1080/03055698.2020.1833840

Lades, L. K., Laffan, K., Daly, M., & Delaney, L. (2020). Daily emotional wellLbeing during the COVIDL19 pandemic. British journal of health psychology, 25(4), 902–911.

Lahaie, C., Earle, A., & Heymann, J. (2013). An uneven burden: Social disparities in adult caregiving responsibilities, working conditions, and caregiver outcomes. Research on Aging, 35(3), 243–274.

Lieu C. A teacher’s diary of a week of school closure - education week. Teacher. Published online March 25, 2020. Accessed May 8, 2020. https://www.edweek.org/tm/articles/2020/03/25/a-teachers-diary-of-a-week-of.html

Meltzer, H., Ford, T., Goodman, R., & Vostanis, P. (2011). The burden of caring for children with emotional or conduct disorders. International Journal of Family Medicine, 2011.

Müller, L. M., & Goldenberg, G. (2020). Education in times of crisis: The potential implications of school closures for teachers and students. A review of research evidence on school closures and international approaches to education during the COVID-19 pandemic. Chartered College of Teaching.

Oberle, E., & Schonert-Reichl, K. A. (2016). Stress contagion in the classroom? The link between classroom teacher burnout and morning cortisol in elementary school students. Social Science & Medicine, 159, 30–37.

Palmer, M., Rose, D., Sanders, M., & Randle, F. (2012). Conflict between work and family among New Zealand teachers with dependent children. Teaching and Teacher Education, 28(7), 1049–1058.

Patrick, S. W., Henkhaus, L. E., Zickafoose, J. S., Lovell, K., Halvorson, A., Loch, S., … & Davis, M. M. (2020). Well-being of parents and children during the COVID-19 pandemic: a national survey. Pediatrics, 146(4). https://doi.org/10.1542/peds.2020-016824

Pinquart, M., & Sörensen, S. (2003). Associations of stressors and uplifts of caregiving with caregiver burden and depressive mood: a meta-analysis. The Journals of Gerontology Series B: Psychological Sciences and Social Sciences, 58(2), 112–128.

Power, K. (2020). The COVID-19 pandemic has increased the care burden of women and families. Sustainability: Science, Practice and Policy, 16(1), 67–73.

Radloff, L. S. (1977). The CES-D scale: A self-report depression scale for research in the general population. Applied psychological measurement, 1(3), 385–401.

Rodrigues, G (March 17, 2020). Ontario government declares state of emergency amid coronavirus pandemic”. Global News. Retrieved Sept 19, 2021.

Rosenzweig, J. M., Brennan, E. M., Huffstutter, K., & Bradley, J. R. (2008). Child care and employed parents of children with emotional or behavioral disorders. Journal of Emotional and Behavioral Disorders, 16(2), 78–89.

Russell, B. S., Hutchison, M., Tambling, R., Tomkunas, A. J., & Horton, A. L. (2020). Initial challenges of caregiving during COVID-19: caregiver burden, mental health, and the parent–child relationship. Child Psychiatry & Human Development, 51(5), 671–682.

Sales, E., Greeno, C., Shear, M. K., & Anderson, C. (2004). Maternal caregiving strain as a mediator in the relationship between child and mother mental health problems. Social Work Research, 28(4), 211–223.

Sokal, L., Trudel, L. E., & Babb, J. (2020). Canadian teachers’ attitudes toward change, efficacy, and burnout during the COVID-19 pandemic. International Journal of Educational Research Open, 1, 100016. https://doi.org/10.1016/j.ijedro.2020.100016

Spitzer, R.L., Kroenke, K., Williams, J.B.W., Löwe, B. (2006). A brief measure for assessing Generalized Anxiety Disorder: The GAD-7. Arch Intern Med, 66(10):1092–1097.

The Kindergarten Program (2016). Retrieved from: https://files.ontario.ca/books/edu_the_kindergarten_program_english_aoda_web_oct7.pdf

Turner C, Adame D, Nadworny E. “There’s A Huge Disparity”: What Teaching Looks Like During Coronavirus. http://NPR.org. Published online 2020. Accessed May 8, 2020. https://www.npr.org/2020/04/11/830856140/teaching-without-schools-grief-then-a-free-for-all

Wang, G., Zhang, Y., Zhao, J., Zhang, J., & Jiang, F. (2020). Mitigate the effects of home confinement on children during the COVID-19 outbreak. Lancet, 395(10228), 945–947.

Weisberger, M., Grinshtain, Y., & Blau, I. (2021). How do technological changes in formal education shape the social roles of teachers who are mothers? Teaching and Teacher Education, 103, 103344.

Whitehead, B. R., & Torossian, E. (2021). Older adults’ experience of the COVID-19 pandemic: A mixed-methods analysis of stresses and joys. The Gerontologist, 61(1), 36–47.

Wolters, C. A., & Daugherty, S. G. (2007). Goal structures and teachers’ sense of efficacy: Their relation and association to teaching experience and academic level. Journal of Educational Psychology, 99(1), 181–193.

Wu, M., Xu, W., Yao, Y., Zhang, L., Guo, L., Fan, J., & Chen, J. (2020). Mental health status of students’ parents during COVID-19 pandemic and its influence factors. General Psychiatry, 33(4). doi: 10.1136/gpsych-2020-100250

Varner, C. (2020). Parents on the front lines of COVID-19 face tough choices. Canadian Medical Association Journal, 192, 467–468.

Yamamura, E., & Tsustsui, Y. (2021). The impact of closing schools on working from home during the COVID-19 pandemic: evidence using panel data from Japan. Review of Economics of the Household, 1–20. 2101.08476

